# Quantifying the relationship between SARS-CoV-2 wastewater concentrations and building-level COVID-19 prevalence at an isolation residence using a passive sampling approach

**DOI:** 10.1101/2022.04.07.22273534

**Authors:** Patrick T. Acer, Lauren M. Kelly, Andrew A. Lover, Caitlyn S. Butler

## Abstract

SARS-CoV-2 RNA can be detected in the excreta of individuals with COVID-19 and has demonstrated a positive correlation with various clinical parameters. Consequently, wastewater-based epidemiology (WBE) approaches have been implemented globally as a public health surveillance tool to monitor the community-level prevalence of infections. Over 270 higher education campuses monitor wastewater for SARS-CoV-2, with most gathering either composite samples via automatic samplers (autosamplers) or grab samples. However, autosamplers are expensive and challenging to manage with seasonal variability, while grab samples are particularly susceptible to temporal variation when sampling sewage directly from complex matrices outside residential buildings. Prior studies have demonstrated encouraging results utilizing passive sampling swabs. Such methods can offer affordable, practical, and scalable alternatives to traditional methods while maintaining a reproducible SARS-CoV-2 signal. In this regard, we deployed tampons as passive samplers outside of a COVID-19 isolation unit (a segregated residence hall) at a university campus from February 1, 2021 – May 21, 2021. Samples were collected several times weekly and remained within the sewer for a minimum of 24 hours (n = 64). SARS-CoV-2 RNA was quantified using reverse transcription-quantitative polymerase chain reaction (RT-qPCR) targeting the viral N1 and N2 gene fragments. We quantified the mean viral load captured per individual and the association between the daily viral load and total persons, adjusting for covariates using multivariable models to provide a baseline estimate of viral shedding. Samples were processed through two distinct laboratory pipelines on campus, yielding highly correlated N2 concentrations. Data obtained here highlight the success of passive sampling utilizing tampons to capture SARS-CoV-2 in wastewater coming from a COVID-19 isolation residence, indicating that this method can help inform public health responses in a range of settings.

**Highlights:**

- Daily SARS-CoV-2 RNA loads in building-level wastewater were positively associated with the total number of COVID-19 positive individuals in the residence
- The variation in individual fecal shedding rates of SARS-CoV-2 extended four orders of magnitude
- Wastewater sample replicates were highly correlated using distinct processing pipelines in two independent laboratories
- While the isolation residence was occupied, SARS-CoV-2 RNA was detected in all passive samples

## 1. Introduction

Severe acute respiratory syndrome coronavirus 2 (SARS-CoV-2) is the seventh coronavirus known to infect human beings and the causative agent for the COVID-19 pandemic (Andersen et al., 2020; Nalbandian et al., 2021). While COVID-19 is most notably known for causing respiratory illness, more recently, it is being recognized as a multi-organ disease with a broad range of extrapulmonary manifestations, including gastrointestinal symptoms (Gupta et al., 2020). The pooled prevalence of gastrointestinal symptoms in COVID-19 patients is about 15%, with diarrhea, loss of appetite, and nausea/vomiting being the most common symptoms (Ren et al., 2020). The mean duration of SARS-CoV-2 RNA shedding in stool is estimated to be roughly 17 days (Cevik et al., 2021). However, prolonged fecal shedding of SARS-CoV-2 RNA has been documented up to 47 days following symptom onset, sometimes persisting in stool for more than two weeks after negative oropharyngeal swabs (Cheung et al., 2020; Fei Xiao et al., 2020; Ling et al., 2020; Y. Wu et al., 2020; Cevik et al., 2021; Nalbandian et al., 2021). It has been estimated that SARS-CoV-2 RNA in stool is detected in nearly 66% of infected patients and can be measured in feces as soon as two days after symptom onset (Lo et al., 2020; Chan et al., 2021). Evidence suggests that both symptomatic and asymptomatic individuals shed SARS-CoV-2 in both respiratory and stool specimens, highlighting the critical role WBE can play in capturing asymptomatic or subclinical cases that may otherwise not be identified (Wu et al., 2020; Xiao et al., 2020; Han et al., 2020; Lo et al., 2020; Wölfel et al., 2020; Zhang et al., 2020; Zhou et al., 2020). Yet, differences in viral shedding rates based on symptom status and disease time course have been detailed, making it difficult to quantitatively interpret SARS-CoV-2 wastewater concentrations, especially to estimate infection prevalence (Zheng et al., 2020). Moreover, additional predictor variables capturing community infection dynamics, disease progression, temporal variations, and fecal shedding kinetics are likely important to consider if possible when estimating caseloads based on SARS-CoV-2 wastewater signatures (Mcmahan et al., 2020; Karthikeyan, et al., 2021; Miura et al., 2021; Proverbio et al., 2021).

Many higher educational institutions and municipalities globally have recognized the utility of wastewater-based epidemiology (WBE) as a surveillance tool for monitoring infection trends and identifying potential case clusters promptly. Several studies have demonstrated a positive correlation between SARS-CoV-2 RNA concentrations in influent wastewater/solids and COVID-19 case metrics, demonstrating the utility of this approach in complementing clinical reporting, particularly in settings where distinct buildings can be isolated (Peccia et al., 2020; D’aoust et al., 2021; Feng et al., 2021). Additionally, in the absence of widespread clinical testing, WBE can act as a passive health surveillance tool to monitor viral dynamics, as it has been shown to presage new case and subsequent hospitalization rates (Peccia et al., 2020; Karthikeyan et al., 2021). Accordingly, numerous higher educational institutions have adopted building-level wastewater testing in combination with contact tracing and isolation/quarantine protocols to support campus public health and guide clinical testing resources to “hotspot” locations (Barich and Slonczewski, 2021; Betancourt et al., 2021; Gibas et al., 2021; Karthikeyan et al., 2021; Reeves et al., 2021; Travis et al., 2021). Due to the population-level and non-intrusive nature of WBE, it will continue to serve as a valuable means to monitor and inform public health programming and response.

Sewage surveillance is ideally suited to complement widespread clinical testing campaigns. Within WBE, sample collection strategies vary with three predominant methodologies emerging: composite sampling, grab sampling and passive sampling. Autosamplers can mechanically pull wastewater samples routinely with defined time or flow intervals to create representative composite aliquots; however, these instruments are expensive and challenging to maintain in certain environments (Bivins et al., 2021). Grab samples are generally an operationally straightforward option, collecting a volume of wastewater at a single timepoint. However, such samples are susceptible to fecal shedding and flow fluctuations, particularly outside buildings with limited drainage areas, and may provide a biased sample of the wastewater stream (Bivins et al., 2021; Rafiee et al., 2021). Several universities have trialed the use of passive samplers in place of traditional composite or grab sampling methods due to additional cost-effectiveness and ease of deployment (Bivins et al., 2021; Brizee, 2021; Corchis-Scott et al., 2021; Severance, 2021). Schang et al. demonstrated that the concentration of SARS-CoV-2 RNA recovered from passive samplers was positively correlated with the viral wastewater concentrations obtained using traditional sampling methods in communities with low caseloads (Schang et al., 2021). Habtewold et al. demonstrated linear uptake of SARS-CoV-2 virus utilizing gauze and membrane passive devices when deployed between 4 and 48 hours, indicating that passive sampling methods can accumulate SARS-CoV-2 viral fragments over time (Habtewold et al., 2022). In addition, Rafiee et al. demonstrated that Moore swabs (gauze pads with string) performed as well as composite samples over 16 hours and were more sensitive than grab samples (Rafiee et al., 2021). A passive sampling approach may be the only practical option for capturing a representative wastewater sample in resource-constrained settings. However, questions still remain on the viral loading dynamics (sorption-desorption rates) with such sampling methods. The passive sampling technique has demonstrated its utility in qualitatively assessing the presence of SARS-CoV-2 titers in wastewater. Still, rigorous evaluation of the relationship between passive probe samples and epidemiological metrics is urgently needed to ensure optimal use of limited health resources.

To address this gap, we provide the results of 64 independent wastewater samples obtained between February 1, 2021 – May 21, 2021, from a COVID-19 isolation residence. All samples were collected through a passive sampling method using tampons that were placed in the sewer for at least 24 hours. The sampling site was isolated in the sewer shed such that the only waste stream inputs came from infected individuals within the SARS-CoV-2 isolation unit. The primary aims of this study were to: (i) demonstrate that a passive sampling approach yields consistent positive SARS-CoV-2 signals coming from building-level wastewater monitoring with a known number of infected persons, (ii) describe the variability in individual fecal shedding rates quantified through this sampling method, (iii) investigate the associations between the SARS-CoV-2 RNA concentrations extracted from the probes and the corresponding caseload in the building, (iv) compare two independent methods for concentrating wastewater samples and subsequent analysis.

## 2. Materials and methods

### 2.1 Study site

During the course of this study, wastewater samples were collected at an access manhole location roughly 190 feet from a COVID-19 isolation residence. The isolation residence served as a temporary living space for students who tested positive for COVID-19. On-campus students relocated to this space after receiving positive test results and were instructed to remain in isolation for 14 days. The wastewater influent at this sampling location was restricted to the isolation building such that the only inputs in the sewer system came from either infected individuals residing within the building or potentially a small number of staff supporting the students in isolation. Sample collection occurred throughout the Spring 2021 academic semester; samplers were deployed for a minimum of 24 hours. Nearly 90% of samples were deployed for roughly 24 hours; however, a minority were deployed for extended periods (weekend). Three to four separate sampling rounds occurred weekly from February 1, 2021 – May 21, 2021. Immediately following collection, all samples were stored at +4 °C until further processing.

### 2.2 Passive swabs and wastewater processing

We utilized tampons made from two types of rayon with a polyester string (OB Applicator Free Tampons, Ultra Absorbency). Two tampons were tied onto separate lengths of 1/8-inch nylon paracord and deployed concurrently at the collection site. After 24 hours, both tampons were collected and transferred to sterile 500mL high-density polyethylene (HDPE) wide-mouth bottles (ThermoFisher Scientific, Waltham, MA, USA). Both tampons were mixed with a total of 200mL of Milli-Q water, vortexed for 1 minute at 3,200 RPM, and pressed utilizing a citrus squeezer to remove maximum liquid from swabs. The volume obtained from each sampling event ranged from 52mL to 80mL, resulting in a total sample volume ranging from 252mL to 280mL after the addition of Milli-Q water. Bovine Respiratory Syncytial Virus (BRSV) was spiked into each sample in a 1:10,000 volume ratio resulting in 1,000 BRSV gene copies per mL in the wastewater sample matrix (Inforce 3 Cattle Vaccine, Zoetis). BRSV was introduced as a surrogate for SARS-CoV-2, functioning as a matrix recovery control for all samples. Following the BRSV spike, samples were homogenized using a vortex mixer (3,200 RPM) and left to incubate at room temperature (RT) for 30 minutes. Samples were then immediately frozen at -80 °C for future processing.

### 2.3 Protocol 1: SARS-CoV-2 concentration and quantification

In June 2021, all samples were removed from -80 °C and thawed over five days at +4 °C prior to processing. Once thawed, samples were centrifuged at 5,500 RPM in 250mL HDPE bottles for 10 minutes at +4 °C. The sample supernatant was decanted into sterile 250mL HDPE bottles for filtration; the remaining pellet was discarded. We utilized a vacuum filtration assembly and generally followed the concentration procedure outlined in detail by Bivins et al. in their protocol (https://dx.doi.org/10.17504/protocols.io.bhiuj4ew). Briefly, we used one vacuum filtration assembly and filtered 50mL of the sample through 0.45μM mixed-cellulose ester membrane filters in duplicate (Pall Corporation, Port Washington, NY, USA). Following filtration, the filter was halved and transferred aseptically into a 0.70mm garnet bead tube in preparation for RNA extraction (Qiagen, Germantown, MD, USA).

Before extraction, 500μL of Buffer AVL (Qiagen, Germantown, MD, USA) and 6.5μL of β-mercaptoethanol (MP Biomedicals, Irvine, CA, USA) were added to the garnet bead tubes. Tubes were bead beat for 60 seconds at 3,200 RPM for four cycles and centrifuged for 30 seconds between each homogenization cycle at 5,000 x g. After completion of bead beating, tubes were centrifuged for a final time at 16,000 x g for 3 minutes, and 140μL of sample was transferred into a 1.5mL microcentrifuge tube in preparation for extraction. RNA was then extracted using Qiagen’s protocol utilizing the QIAamp Viral RNA Mini Kit (Qiagen, Germantown, MD, USA). Purified RNA concentrate was eluted with 80μL of Buffer AVE (Qiagen, Germantown, MD, USA) and held at +4 °C briefly. Total RNA in each sample was measured using an RNA high sensitivity assay kit on the Qubit 4.0 prior to RT-qPCR quantification (ThermoFisher Scientific, Waltham, MA, USA).

SARS-CoV-2 was quantified using real-time qPCR, applying SYBR chemistry (Luna^®^ Universal One-Step qPCR Master Mix, Cat No. E3005E). The N1 and N2 genes unique to SARS-CoV-2 were quantified using the 2019-nCoV N1 forward (Cat No. 10006830) and reverse (Cat No. 10006831) primers and the 2019-nCoV N2 forward (Cat No. 10006833) and reverse (Cat No. 10006824) primers synthesized by Integrated DNA Technologies (IDT). Standard curves for the N1 and N2 analyses were generated by quantifying a synthesized SARS-CoV-2 plasmid manufactured by IDT (Cat No. 10006625). The standard was assayed in triplicate following a 10-fold serial dilution from 2,000 gene copies/μL to 0.2 gene copies/μL. All RNA samples were assayed in triplicate. Samples were processed in biological replicates, generating six RT-qPCR data points per individual sample. Each 20μL reaction mix contained the following: 4μL of template RNA; 10μL of 2X Luna Universal One-Step Reaction Mix; 1μL of 20X Luna WarmStart^®^ RT Enzyme Mix; 1.6μL of primers; and 3.4μL of PCR-grade water. PCR analysis was conducted using a BioRad 96-well real-time PCR system, with the following cycle parameters: 55 °C for 10 min; 95 °C for 1 min; 40 cycles x (95 °C for 10 sec, 62 °C for 30 sec); 60-95 °C in 5 second increasing increments of 0.5 °C. Melt curves were analyzed to ensure the amplification of a single target PCR amplicon. No-template controls (NTCs) were assayed in triplicate for all PCR runs for quality control. Following successful sample quantification, all wastewater samples were again stored at -80 °C. Standard curves were used to quantify N1 and N2 gene copies in the polymerase chain reaction, which were converted to gene copies/L of raw wastewater captured by our passive samplers (Supplemental Document). All samples that failed to meet the following criteria were re-assayed: i) standard curves with R^2^ > 0.985, ii) primer efficiency between 90%-120%, iii) no signs of PCR inhibition or non-specific amplification. The y-intercept value for all runs ranged from (35.1, 36.6), and the slope values ranged from (−3.33, - 2.93). The level of detection (LOD) for this assay was estimated to be 1.6 × 10^4^ SARS-CoV-2 gene copies/L of wastewater. BRSV quantification occurred using an identical RT-qPCR procedure but with custom BRSV primers (ThermoFisher Scientific, Waltham, MA, USA).

### 2.4 Protocol 2: SARS-CoV-2 concentration and quantification

In October 2021, all 64 raw wastewater samples were removed from -80 °C and thawed over five days at +4 °C. These samples were quantified for a second time using a unique concentration method through a separate RNA extraction and RT-qPCR pipeline. We followed an adapted version of the SARS-CoV-2 Wastewater RNA Concentration Protocol written at the University of Connecticut (https://dx.doi.org/10.17504/protocols.io.bn58mg9w). Once raw wastewater samples were thawed, 40 mL of sample was added to a 50mL conical tube. 600μL of affinity-capture magnetic hydrogel nanoparticles in solution at a 5 mg/mL concentration was spiked into the sample (Ceres Nanosciences, Manassas, VA, USA). Samples were homogenized and incubated at RT for 20 minutes. After incubation, samples were placed on custom-built magnetic racks for 30 minutes at RT for viral concentration. This magnetic separation protocol allowed for the easy removal of supernatant while retaining a pellet of nanoparticles with the entrapped viral fragments. 1.2mL of DNA/RNA Shield (Zymo Research, Irvine CA, USA) was added to each pellet. Samples were briefly homogenized and incubated at 56 °C (ten minutes) to release the viral fragments from the nanoparticle matrices. Samples were returned to the magnetic racks for 10 minutes to separate the sample lysate from the nanoparticles. 500μL of the sample was aliquoted into technical replicate test tubes to be further processed at the Institute for Applied Life Sciences Clinical Testing Center (ICTC), which obtained its state clinical laboratory license (CLIA) in October 2020. Briefly, ICTC utilized an automated RNA extraction platform (Hamilton Company, Reno, NV, USA) using the MagMax Viral/Pathogen II Nucleic Acid Isolation Kit (ThermoFisher Scientific, Waltham, MA) to isolate and purify nucleic acid. SARS-CoV-2 was quantified targeting the N2 gene with real-time qPCR, applying TaqMan chemistry (Luna® Universal Probe One-Step RT-qPCR Kit, E3006E) using a BioRad 384-well real-time PCR system. Data was reported as extracted gene copies/mL from our concentrated samples, which we converted to gene copies/L of wastewater by using the known dilution factor from the original sample.

### 2.5 Isolation residence case data and clinical surveillance

Aggregated and de-identified COVID-19 case data were obtained from administrative records for the isolation residence on campus during the Spring 2021 academic year. These data consisted of the total number of individuals present in the isolation facility, including sex and self-reported symptoms. The public health surveillance program was approved by the Institutional Review Board (#20-258); this research protocol had a separate filing (approval #21-140). Data were processed by HIPAA-trained staff at the Public Health Promotion Center (PHPC) and were acquired on university-administered systems and HIPAA-compliant platforms. During this timeframe of wastewater testing, all on-campus students were required to be tested twice weekly via nasal swab PCR tests. All off-campus students, faculty, and staff that came to campus were required to be tested once weekly during the semester. On-campus students with a positive SARS-CoV-2 test were required to isolate in the designated isolation residence hall. Off-campus students were provided the option to isolate themselves on campus.

### 2.6 Data analysis

Statistical analyses were performed using SAS 9.4 (SAS Institute, Cary, NC, USA), and visualizations were created using RStudio (ver. 1.4.1103) with ggplot2 (ver. 3.3.5). Slopes and y-intercepts from RT-qPCR were used to quantify copies of SARS-CoV-2 in each reaction using the instrument’s recorded Cq value. This SARS-CoV-2 copy number value was then transformed into a gene copies/L of extracted wastewater value through a series of unit conversations and dilution factors based on our wastewater processing methods (Supplementary Document). Building-level volumetric water metering provided 24-hour building-level water use totals for the isolation residence, which were utilized as a proxy for daily wastewater flow. The flow values were converted from cubic feet to liters. Daily viral loads at the isolation residence were estimated by multiplying [(SARS-CoV-2 gene copies/L of wastewater) X (L of wastewater influent/day)] to provide an aggregate SARS-CoV-2 gene copies/day measurement. Wilcoxon-Mann-Whitney (WMW) tests were utilized to test for statistical differences between the N1 and N2 gene copy concentrations due to the paired comparisons and non-normal distribution of these data. This test was also employed to assess statistical differences between N2 gene concentrations quantified through two separate sample processing and analysis pipelines. We employed multivariable negative binomial models to quantify the relationship between the wastewater viral load and the accompanying building caseload, adjusting for significant covariates. We also evaluated the performance of the negative binomial model utilizing McFadden’s pseudo-R^2^ statistic. All tests were two-sided, with α = 0.05 for hypothesis testing.

## 3. Results and discussion

### 3.1 Student characteristics & performance of passive samples

During the Spring 2021 academic term, individuals in the isolation residence ranged from 17–25 years of age with females making up 40.6% of total cases and males making up 59.4% of cases on wastewater collection days (Supplemental Document). During this time, 92.3% of COVID-19 cases reported having at least one illness-related symptom. A global meta-analysis including nearly 30 million individuals undergoing COVID-19 testing found that 40.5% (95% CI, 33.50%–47.50%) of laboratory-confirmed COVID-19 cases were asymptomatic (Ma et al., 2021). Patient symptom reports occurred at multiple timepoints along the care cascade possibly resulting in the misclassification of cases captured prior to symptom onset. In contrast, cases in the isolation residence were asked to report symptoms daily throughout their illness, which may partially explain the discordance between the high proportion of symptomatic individuals in the isolation residence compared with estimates of symptomatic prevalence found in the general population.

Overall, the tampon swabs consistently captured the SARS-CoV-2 molecular signature from the waste stream. Each sample contained liquid from two tampons yielding between 52mL and 80mL of raw sewage. Total RNA mass extracted from each sample ranged from 1.31 ng/μL to 35.5 ng/μL with a mean concentration of 7.0 ng/μL ± 5.14, indicating that our passive swabs captured fluctuating amounts of RNA in the sewer. Over the 16-week study period, all passive samples captured SARS-CoV-2, demonstrating consistency in tampons amassing detectable amounts of virus. The mean N1 Cq value was 32.84 ± 2.35, and the mean N2 Cq value was 32.37 ± 2.57 for all datapoints obtained using Protocol 1. A Wilcoxon rank-sum test showed no evidence of a statistically significant difference between the median N1 and N2 signals within this study (p = 0.69). Additionally, the mean N2 Cq value provided by ICTC on identical raw wastewater samples processed through Protocol 2 was 26.53 ± 2.63. The increased N2 Cq sensitivity observed using Protocol 2 is likely due to a greater proportion of template RNA in the PCR compared with Protocol 1. Overall, the variability of SARS-CoV-2 viral loads across sample replicates was often larger at lower concentrations, sometimes extending over an order of magnitude (Fig. 1). Inconsistencies seen here across biological and technical replicates have previously been reported when quantifying SARS-CoV-2 titers in wastewater (D’aoust et al., 2021; Graham et al., 2021; Pecson et al., 2021). Throughout the study period, the median N2 load was 1.29 × 10^9^ gene copies per day and the median N1 daily load came in slightly lower at 1.04 × 10^9^ gene copies (Table 1). A general decrease in the day-to-day viral load from February to May is apparent throughout the semester (Fig. 1).

**Table 1.**
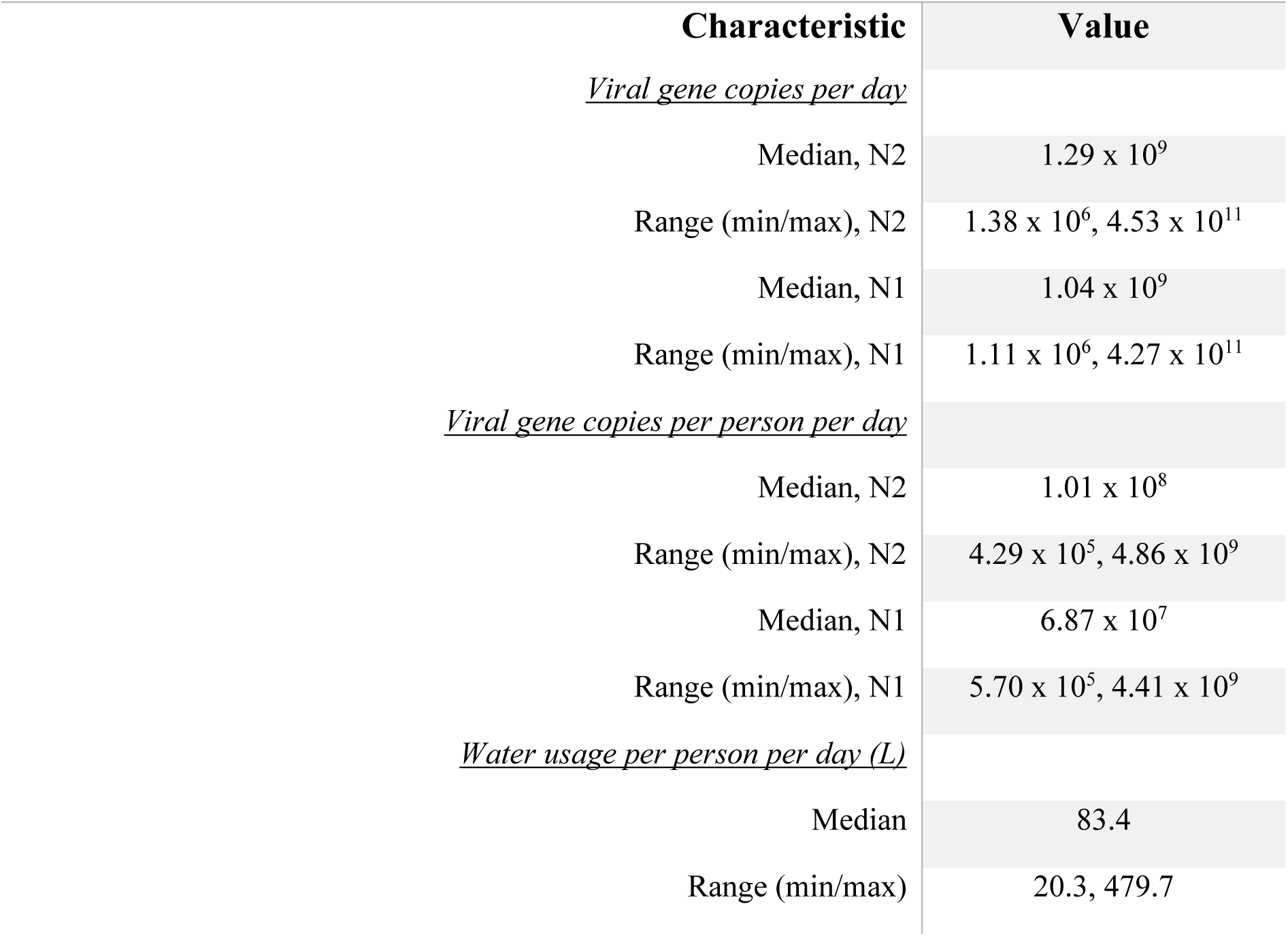
Quantification of viral loads and viral loads per population in the isolation residence

| Characteristic | Value |
| --- | --- |
| <u>*Viral gene copies per day*</u> |  |
| Median, N2 | $1.29 \times 10^9$ |
| Range (min/max), N2 | $1.38 \times 10^6$, $4.53 \times 10^{11}$ |
| Median, N1 | $1.04 \times 10^9$ |
| Range (min/max), N1 | $1.11 \times 10^6$, $4.27 \times 10^{11}$ |
| <u>*Viral gene copies per person per day*</u> |  |
| Median, N2 | $1.01 \times 10^8$ |
| Range (min/max), N2 | $4.29 \times 10^5$, $4.86 \times 10^9$ |
| Median, N1 | $6.87 \times 10^7$ |
| Range (min/max), N1 | $5.70 \times 10^5$, $4.41 \times 10^9$ |
| <u>*Water usage per person per day (L)*</u> |  |
| Median | 83.4 |
| Range (min/max) | 20.3, 479.7 |

**Fig 1.**
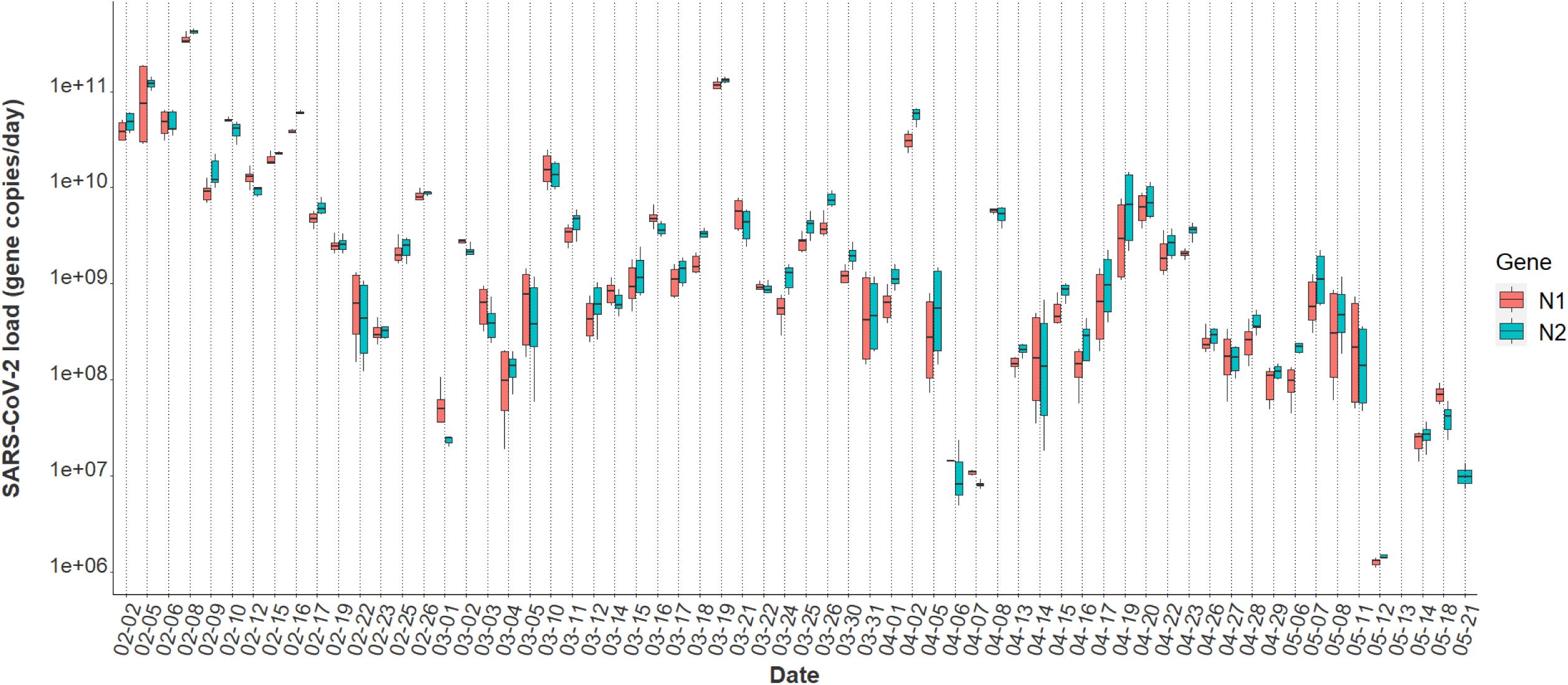
N1 and N2 daily viral loads from the COVID-19 isolation residence, February 1, 2021 – May 21, 2021 (n=64).

### 3.2 Comparison of SARS-CoV-2 detection

A statistically significant correlation between mean N1 and N2 wastewater concentrations was observed throughout the study period (r = 0.96), and comparable longitudinal wastewater trends are apparent (Fig. 1). These data suggest that either gene may be utilized to quantify SARS-CoV-2 to develop trends when using passive probes. Additionally, a strong positive association was noted between independent N2 wastewater concentrations on identical samples processed through distinct concentration, extraction, and RT-qPCR methods (r = 0.87) (Fig. 2). The average raw N2 wastewater concentration quantified in our lab with Protocol 1 was 3,873 ± 10,330 N2 gene copies/mL of wastewater. In comparison, Protocol 2 yielded an average of 3,661 ± 6,883 N2 gene copies/mL of wastewater. A Wilcoxon rank-sum test found no statistically significant difference between the average N2 concentrations quantified through these separate methods (p = 0.17). A high degree of reproducibility in SARS-CoV-2 quantification was evident between the two laboratories suggesting that a standardized method for processing wastewater samples may not be overly critical for obtaining valuable data to support public health decision-making. Similar conclusions were noted when 32 independent laboratories processed replicate wastewater samples from two major wastewater treatment plants in Los Angeles County to quantify SARS-CoV-2 using independent SOPs (Pecson et al., 2021). After correcting for recovery rates, 80% of the SARS-CoV-2 wastewater concentration data fell within a range of approximately ±1log GC/L coming from groups utilizing eight distinct methods (Pecson et al., 2021).

**Fig 2.**
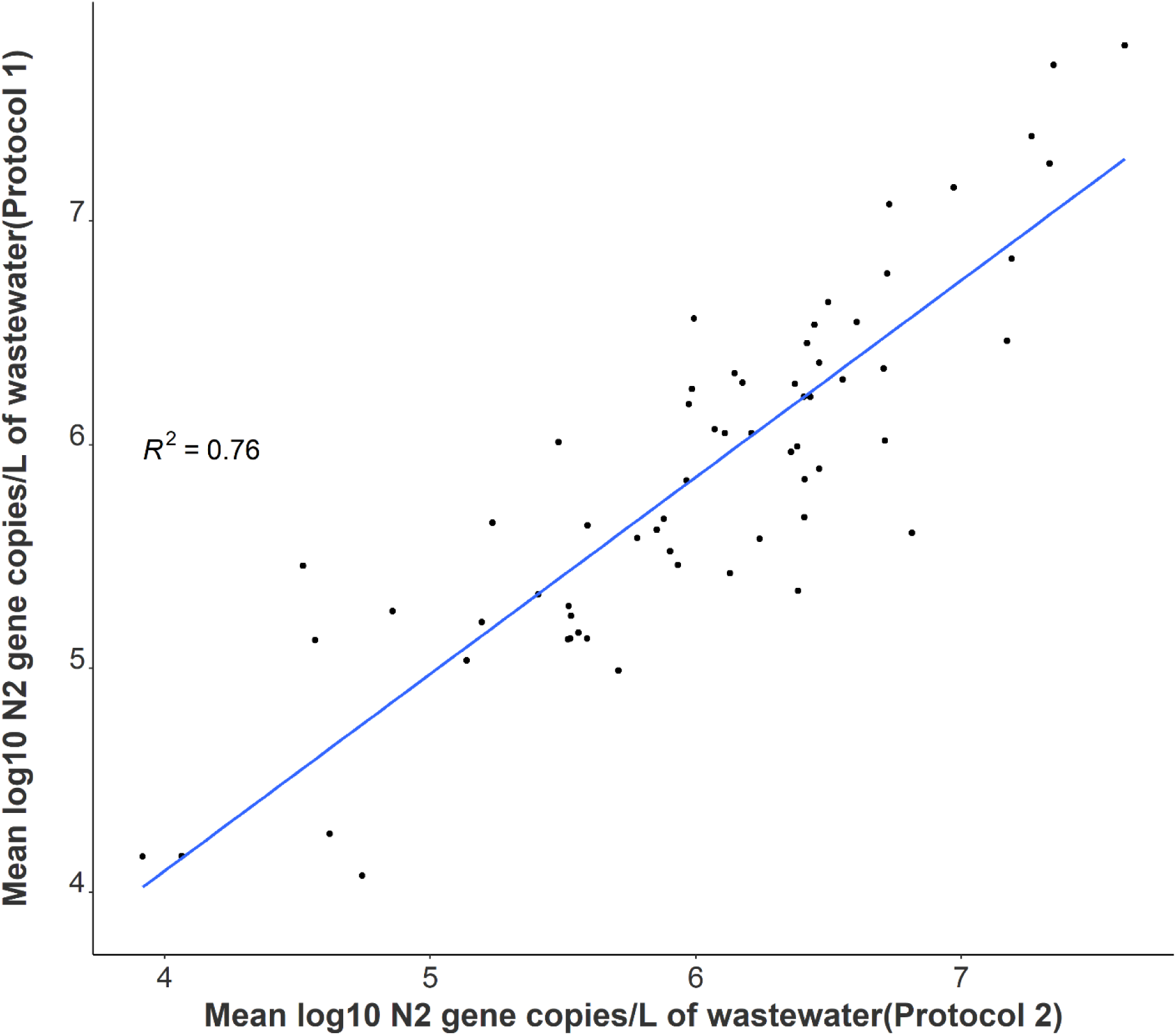
Correlation between independent average N2 log-transformed viral concentrations on identical samples utilizing separate processing and analysis pipelines (n = 64).

### 3.3 Variability in SARS-CoV-2 signal

Variation in daily N1 and N2 viral loads per individual in isolation throughout the study duration extended greater than four orders of magnitude based on results from our passive swabs (Fig. 3). The median N2 load throughout the study was measured at 1.01 × 10^8^ gene copies per individual per day while the median N1 load per individual was 6.87 × 10^7^ gene copies per day (Table 1). The observed variation in fecal shedding rates is consistent with several studies reporting levels of SARS-CoV-2 in stool ranging from 5.5 × 10^2^ − 1 × 10^7^ gene copies/mL, which translates to 5.3 × 10^7^ − 9.61 × 10^11^ expected daily gene copies per infected individual in isolation when accounting for the mean individual daily building-level water use production (96 liters) at the isolation building (Han et al., 2020; Pan et al., 2020; Wölfel et al., 2020; Zhang et al., 2020). Since we could not obtain data on the temporal complexities of disease progression for each infected individual, fecal shedding rates were assumed constant for the above calculation. Differences in building-level water use indicate significant variation in personal hygiene habits and bathroom behaviors between students (Fig. 4). Therefore, substantial variation in individual viral loads per day was expected in this study due to discrepancies in viral shedding, temporal disease dynamics, and water use behavior.

**Fig 3.**
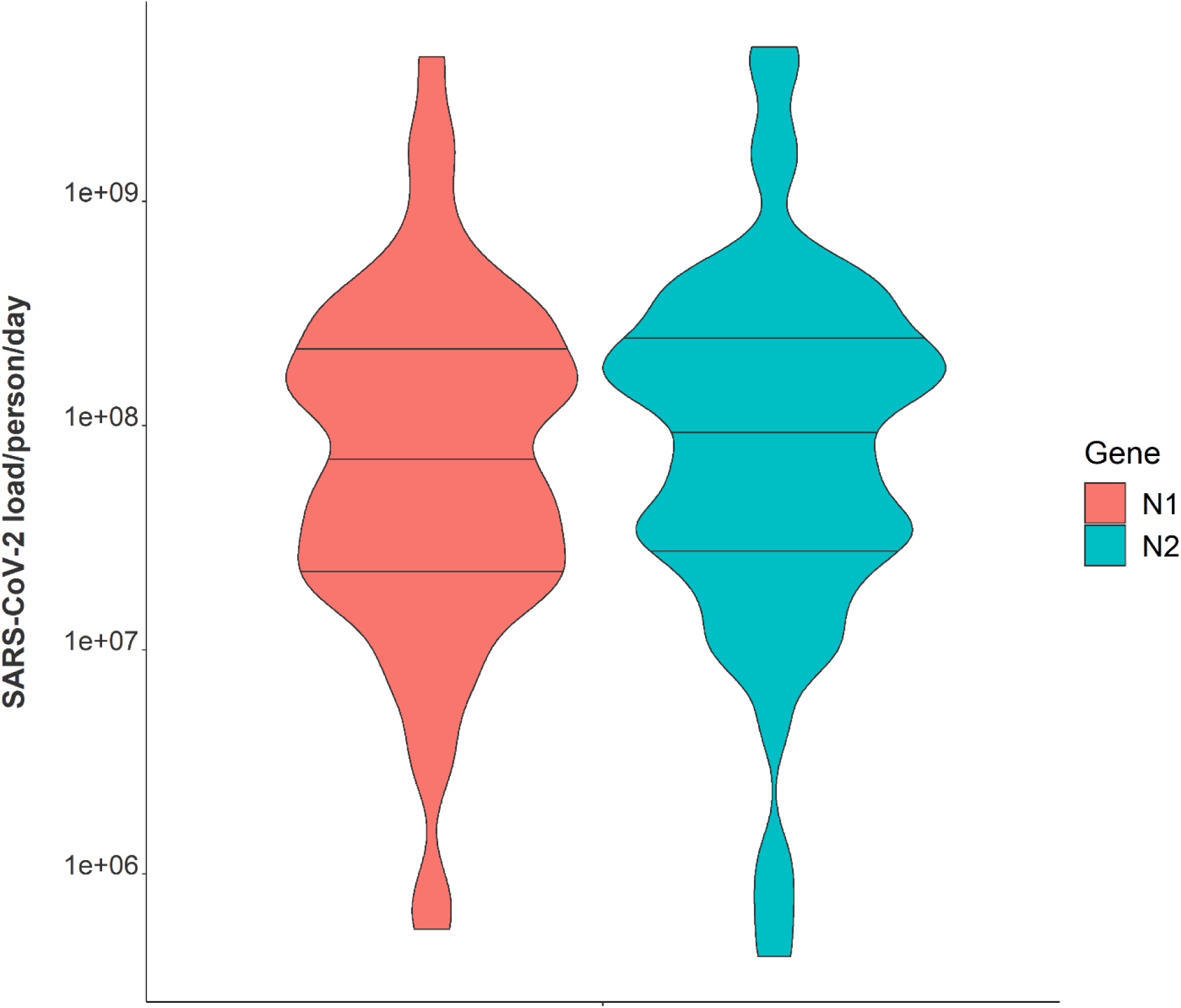
Violin plot showing distributions of N1 and N2 average daily wastewater viral loads per individual in isolation from February 1, 2021 – May 21, 2021. Note: Markers shown are median, 25^th^ and 75^th^ quantiles.

**Fig 4.**
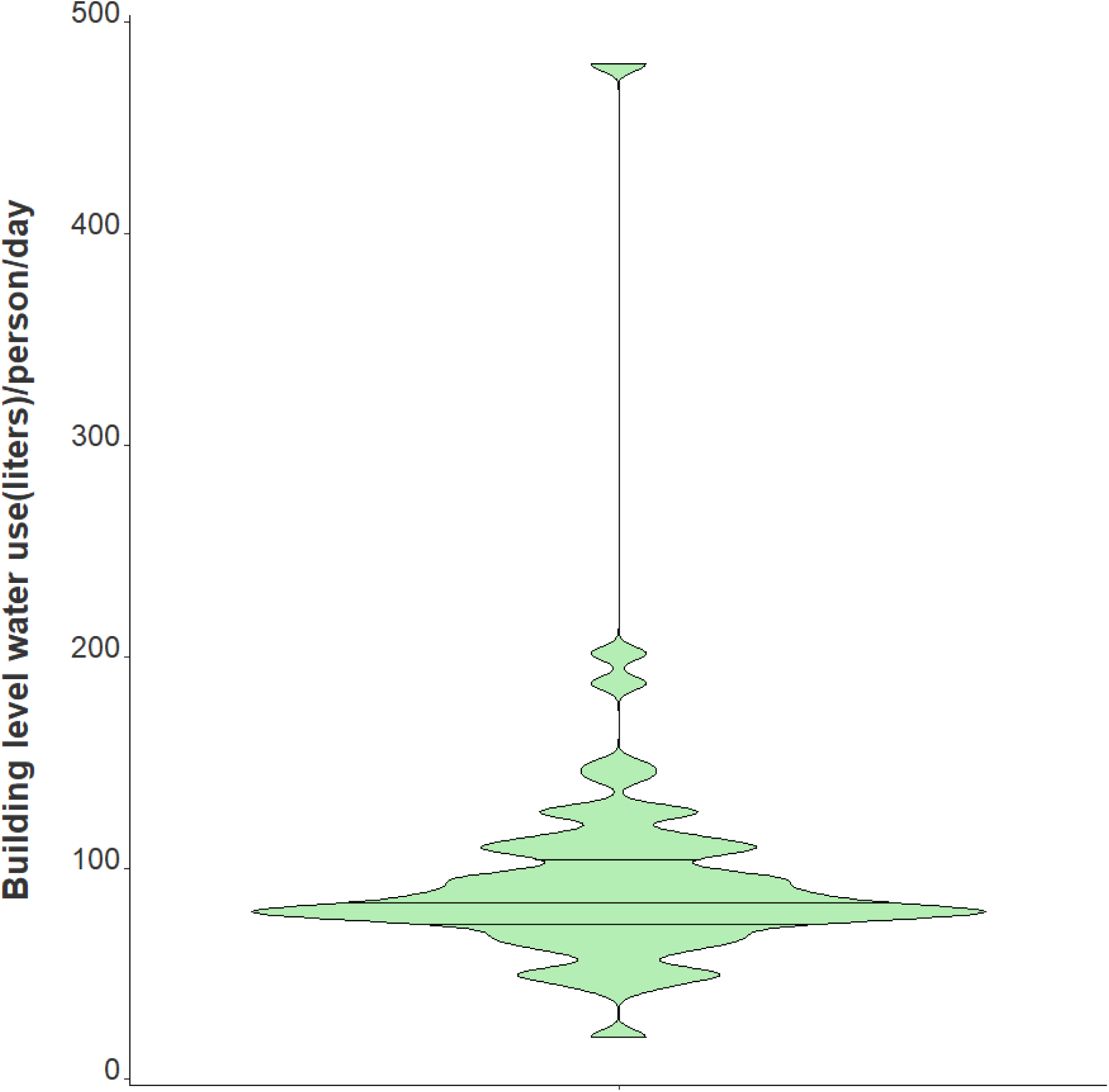
Violin plot showing distribution of average daily building-level water use per individual in isolation from February 1, 2021 – May 21, 2021. Note: Markers shown are median, 25^th^ and 75^th^ quantiles.

Current campus-wide wastewater surveillance activities demonstrate that this passive sampling approach is consistently capturing human RNase P as our fecal content biomarker as well as the N2 gene in those catchments where students have a positive laboratory-based COVID-19 test. Our initial observations across campus sampling indicate that tampons effectively capture SARS-CoV-2 viral particles in diverse sewer catchment areas with fluctuating proportions of non-infected and infected individuals; still, additional work is being conducted to explore the quantitative utility of this approach at unique sampling locations.

### 3.4 Relationships between passive samplers and epidemiological reporting

It has been demonstrated that the quantification of the SARS-CoV-2 virus in wastewater is a useful epidemiological tool to develop and complement longitudinal infection trends when using active sampling approaches (Castro Gutierrez et al., 2021; Graham et al., 2021). However, questions remain on whether passive swabs are limited to providing binary positive/negative SARS-CoV-2 wastewater results, or if there is utility in quantification and correlation to clinical testing metrics over time. The SARS-CoV-2 wastewater load in the isolation residence peaked on February 8, 2021, at 4.53 ×10^11^ N1 gene copies/day, which was coincident with the isolation residence occupancy peak of 222 students (Fig. 5). We evaluated the association between the building occupancy and the 24-hour SARS-CoV-2 load using negative binomial models, adjusting for the mean BRSV recovery (mean ± SD, 14.0% ± 14.1) and the daily percentage of female occupants in the building as these covariates were significantly associated with occupancy (Table 2). The model was evaluated using McFadden’s pseudo R^2^ as a goodness-of-fit measure (Supplemental Document).

**Table 2.**
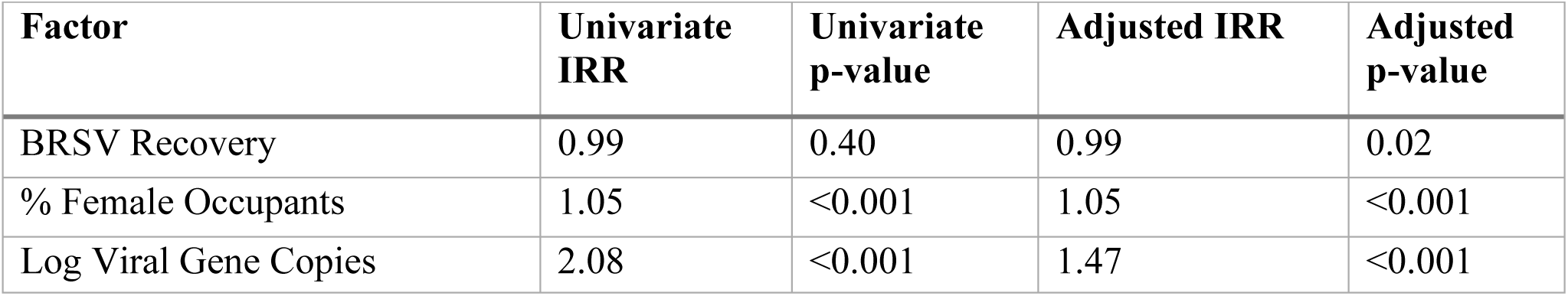
Time series negative binomial models to quantify associations between captured viral loads and defined patient populations, Massachusetts 2021. Note: IRR = Incidence Rate Ratio.

**Fig 5.**
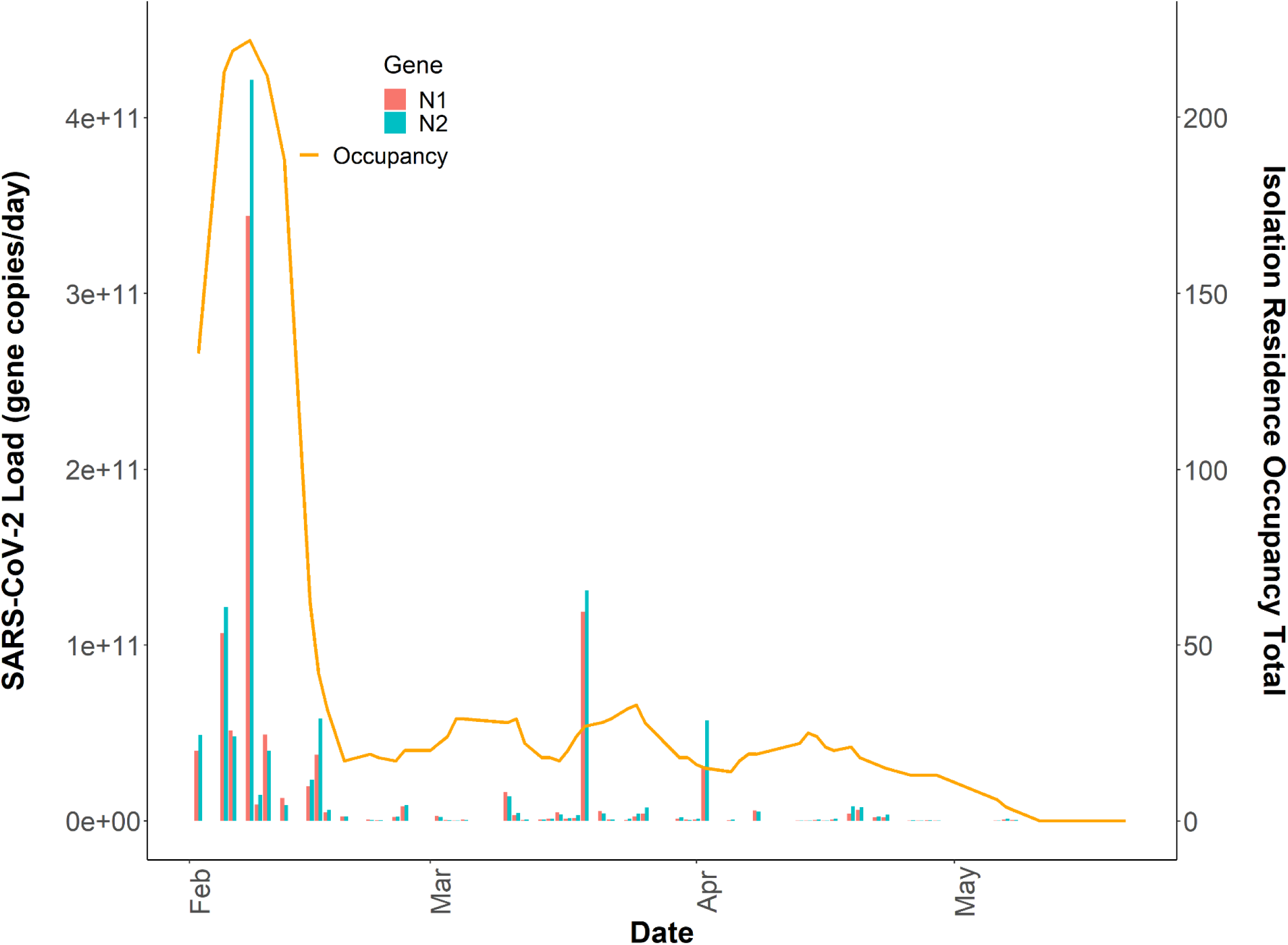
Total daily isolation building occupancy (line) plotted with N1 and N2 gene copies/day (bars) during the Spring 2021 academic semester. Note: Both SARS-CoV-2 daily wastewater viral loads and daily isolation residence occupancy totals are reported on linear y-axes (n=64).

A one unit increase in the log-transformed wastewater SARS-CoV-2 daily load was associated with a 47% increase in the total building occupancy, adjusting for BRSV recovery and the percentage of females in the building (Incidence Rate Ratio (IRR)= 1.47 (95% CI: 1.27–1.71, p<0.001). After adjustment for BRSV recovery and the daily SARS-CoV-2 load, we found a 5% increase in occupancy count for each one percentage increase in females occupying the isolation residence (IRR = 1.05; 95% CI: 1.04–1.06, p<0.001). Several factors may have contributed to this observation, including but not limited to differences in fecal shedding intensity between females and males as a result of disease severity or time course of illness. A meta-analysis on SARS-CoV-2 RNA fecal shedding revealed that patients with gastrointestinal (GI) symptoms had a 2.4-fold increased likelihood of excreting detectable levels of SARS-CoV-2 RNA in their stool compared with those with no GI symptoms (Odds Ratio (OR)= 2.4, 95% CI: 1.2–4.7) (Zhang et al., 2021). Moreover, a cross-sectional study conducted in Poland examining sex differences in COVID-19 symptoms found that self-reported gastrointestinal symptoms among non-hospitalized patients were significantly higher in females than men (Sierpiński et al., 2020). Together, these suggest that on average, a greater proportion of females in the isolation residence were shedding detectable levels of virus compared to men as a result of sex-linked differences in the prevalence of GI symptoms from COVID-19. We used our final model to predict the Spring 2021 occupancy in the isolation residence and compared these results with actual case numbers. The results suggest that utilizing tampons for passive sampling in wastewater is a viable option to predict building-level caseloads (Fig. 6).

**Fig 6.**
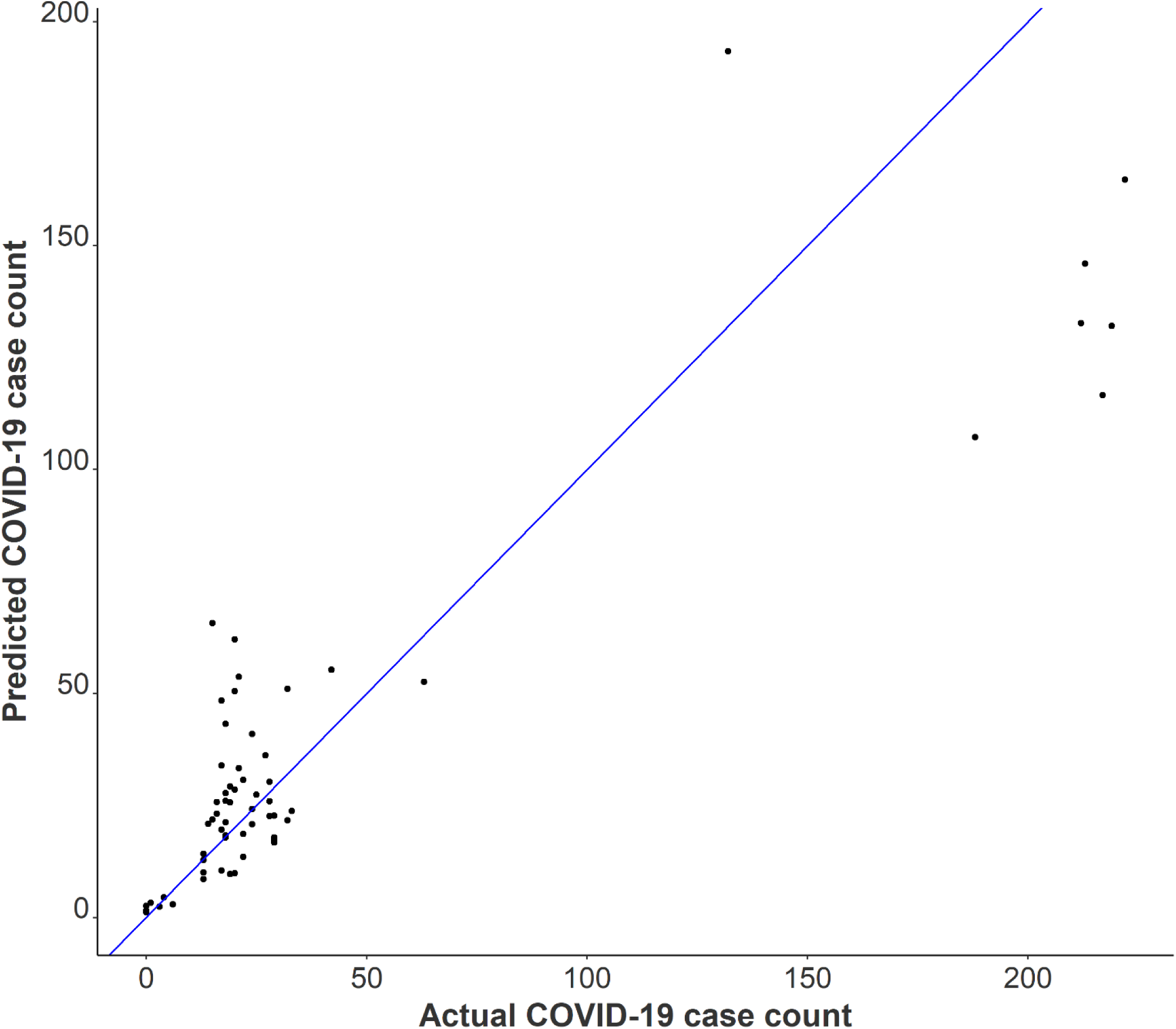
Observed COVID-19 isolation residence occupancy plotted against predicted COVID-19 isolation residence occupancy using negative binomial modeling with Spring 2021 data.

It is possible that incorporating additional covariates such as time since symptoms or diagnosis, sample fecal load, or obtaining more time-resolved flow data could help improve the model by capturing additional changes associated with viral shedding dynamics and SARS-CoV-2 loading onto passive samplers. Additionally, it is important to note that we captured several detectable SARS-CoV-2 signals in the sewer spanning ten days (May 11, 2021–May 21, 2021) after the last individual had exited the isolation residence (Fig. 1). This observation strongly suggests that viral fragments can remain in sewer systems for an extended period despite no active cases in a catchment and that SARS-CoV-2 decay in low-flow environments can occur over a timeframe of weeks. It has been demonstrated that SARS-CoV-2 RNA can accumulate in sewer biofilms which could have served as the primary source of viral RNA during this timeframe (Morales Medina et al., 2021). SARS-CoV-2 persistence in the sewer may have important implications for signal interpretation as positive cases move from their regular living quarters to isolation on college campuses or as students return to college campuses after break periods.

### 3.5 Limitations

Several characteristics relating to this research need to be further explored, and various limitations should be considered in generalizing the findings outlined above. First, most passive samples (nearly 90%) remained in the sewer for 24 hours; however, some were left at the isolation residence for more than one day. A sensitivity analysis excluding these samples left in the sewer for an extended period shows a 44% increase in the occupancy count for each one unit increase in the log-transformed wastewater SARS-CoV-2 daily load, adjusting for BRSV recovery and the percentage of females in the building (IRR 1.44, 95% CI 1.22-1.70). Though only students with a positive COVID-19 clinical test resided in the isolation building, we cannot exclude the possibility of staff members contributing to the building-level water use, which could bias results. Also, it is necessary to note that the quantification of SARS-CoV-2 gene copies/L of wastewater, as measured using the raw influent sewage captured by our passive samplers, may not precisely represent the actual composition of sewage throughout the 24-hour sampling period. Instead, our quantification of SARS-CoV-2 gene copies in the wastewater comes from the “extracted” wastewater over the 24-hour time span, which was required to normalize the N1 and N2 signals to daily flow conveniently. Moreover, hourly flow data and bathroom-level flush counts may have provided more information on day-to-day student behavior. All samples were frozen at -80 °C for differing periods of time prior to quantification which could have resulted in differential SARS-CoV-2 signal degradation. Lastly, incorporating normalization parameters to account for changes in the daily wastewater contributions could be important to include in future models (e.g., human fecal normalization marker). Still, it remains unclear if the addition of other covariates could better elucidate complex viral shedding dynamics to accurately estimate building-level caseloads. Even so, data presented here indicate that the duration and direction of trend classification is a practicable surveillance application using a passive sampling approach. Our data indicate a clear relationship between daily viral wastewater concentrations and building-level infection prevalence. We provide a proof of concept that increased SARS-CoV-2 concentrations in sewage, indicated by a greater number of infected individuals, yield an increased accumulation of viral fragments on our passive samplers over 24 hours (Fig. 4).

## 4. Conclusions

This study investigated the functionality of tampons as passive samplers in capturing SARS-CoV-2 viral fragments in building-level raw sewage. Passive swabs consistently captured virus at a COVID-19 isolation residence over 16 weeks, indicating that this method is a feasible option to identify residential halls with infected individuals shedding SARS-CoV-2. Data here also shed light on the quantitative potential of the captured daily wastewater viral load with 24-hour passive probes and its relation to building-level COVID-19 prevalence as measured via self-administered nasal swabs. The positive association found between the daily viral wastewater load and the isolation building occupancy demonstrates the ability of passive samplers to capture increased SARS-CoV-2 concentrations in influent sewage. Again, additional normalization parameters, including human fecal indicators, could be explored to illustrate building-level COVID-19 caseloads from the captured SARS-CoV-2 wastewater signal. Furthermore, the considerable variability observed in individual viral fecal shedding rates should be considered in future work if accurately predicting the prevalence of COVID-19 in a residential building is a primary goal.

We demonstrated that SARS-CoV-2 quantification was highly correlated between the N1 and N2 gene and N2 wastewater concentrations measured through different processing pipelines. Here, we provide evidence that universities can use either gene to develop and complement existing COVID-19 trends. We demonstrate that independent wastewater processing and quantification methods provide statistically similar and equally useful wastewater data. The prevailing method for obtaining wastewater samples in WBE involves autosamplers collecting liquid composite samples. However, passive samplers not only ease the burden of deployment efforts and sampling expenses, but such samplers may capture variabilities in the wastewater stream missed by time-weighted autosamplers through continuous exposure to raw influent sewage. Given the sensitivity, low cost, and practicality of deploying tampons as passive samplers coupled with the significant positive correlation observed between the daily wastewater viral load and the caseload in the isolation residence over time, we consider this method to be a valuable public health tool for COVID-19 surveillance. In conclusion, this paper provides evidence that tampons can provide a reproducible and informative SARS-CoV-2 signal from wastewater which is particularly relevant for resource-limited communities interested in conducting and operationalizing building-level COVID-19 wastewater surveillance.

## Supporting information

Manuscript.docx(wouldn't properly convert to pdf)

## Data Availability

All data produced in the present study are available upon reasonable request to the authors

## Credit authorship contribution statement

P. Acer conceptualized the study design, collected the data, conducted laboratory-based analysis, performed data analysis/visualization and was the original draft writer of this manuscript. C. Butler was the supervising principal investigator on this work and supported P. Acer in the design and execution of the experiment and supported the writing, reviewing, and editing of this manuscript. A. Lover supported the collection of public health data and supported data analysis efforts as well as the writing, reviewing, and editing of this manuscript. L. Kelly supported the data acquisition and contributed to the writing and editing of the manuscript.

## Acknowledgements

This research was supported with special funds from the Office of Research & Engagement at the University of Massachusetts and by Mike Malone, Vice Chancellor for Research & Engagement. As well, this research was supported with funds from The Environmental Health Science Summer Research Fellowship supported by the National Institute of Environmental Health Sciences (#R25ES031498). The Institute for Applied Life Sciences Clinical Testing Center was integral in performing the sample analysis reported in this work as part of their clinical testing operations, notably acknowledging Ashley Eaton and Peter Reinhart. This research was also funded by the Massachusetts Department of Transportation Highway Division (MassDOT) under Interagency Service Agreement No. 106059. The views, opinions, and findings contained in this paper are those of its Authors, and do not necessarily reflect the official view or policies of the MassDOT. This paper does not constitute a standard, specification, or regulation.

