## Supplemental Document for "Quantifying the relationship between SARS-CoV-2 wastewater concentrations and building-level COVID-19 prevalence at an isolation residence using a passive sampling approach"

**Wastewater Concentration Calculations**

The Cq value recorded by the PCR machine is first measured against the standard curve of known copies of SARS-CoV-2 genome. The signal is converted from Cq values to the concentration in the PCR using the constants generated by the standard curve for each plate: y-intercept, y_int, and slope (m)

Eqn. 1. $pcr_{conc}=e^{\frac{(CT-y\_int)}{m}} units=\frac{gene copies}{\mu L of PCR mixture}$

Divide by the volume of cDNA template in each well, multiply by the 20 $\mu L$ PCR volume

Eqn. 2. $\frac{gene copies}{\mu L PCR}\times\frac{20 \mu L PCR}{vol. cDNA template}= \frac{gene copies}{\mu L\mathrm{cDNA}}$

The RT reaction prior to PCR was assumed to be 100% efficient, meaning the concentration of cDNA in the reaction is equal to the initial RNA concentration

Eqn.3 ${RT}_{eff}= \frac{conc cDNA}{conc RNA}=1$

Convert to gene copies in the entire RNA stock by multiplying by the total volume of RNA extracted.

Eqn. 4 $\frac{gene copies}{\mu L\mathrm{RNA}} \times V_{extracted}=gene copies$

RNA was extracted from ½ of a filter paper. Multiply by 2 to get the gene copies per whole filter.

Eqn. 5 $gene copies \times2= gene copies$

Divide by the volume of liquid phase sample filtered to obtain the concentration in the **eluted** sample solution.

Eqn. 6 $\frac{gene copies}{vol filtered}=gene copies / L$

Calculate the proportion of sample volume in the eluent, PSE. V_dilute_ is the volume of water added to the sample to help elute particles from the tampon. V_elute_ is the total volume obtained from the tampon sample, including V_dilute_.

Eqn. 7 $PSE= \frac{V_{elute}-V_{dilute}}{V_{elute}} units= \frac{mL}{mL}=[ ]$

Multiply the result from Eqn. 7 to obtain the concentration in the **eluted** sample.

Eqn. 8 $\frac{gene copies}{L} \times PSE=\frac{gene copies}{L}$

Full calculations summarized:

Eqn. 9

$${PCR}_{conc}\times\frac{20 \mu L}{vol. cDNA template}\times V_{extracted}\times\frac{2}{V_{filtered}}\times PSE=\frac{gene copies}{L} of wastewater in eluted sample$$

**Supplemental Table 1.** Demographic and clinical characteristics of isolation building occupants

| Characteristic | Value |
| --- | --- |
| Sex |  |
| Male (%) | 59.4 |
| Female (%) | 40.6 |
| Reported any symptoms (%) | 92.3 |

**Model Evaluation**

To evaluate the negative binomial model, we calculated McFadden’s pseudo R^2^ value as a goodness-of-fit measure, which was found to be 0.20 indicating that our model performs well (McFadden, 1977). This statistic was generated comparing the null model to the fitted model using the following equation:

$$R^{2}=1-\frac{\log\hat{L}\left( M_{Full} \right)}{\log\hat{L}\left( M_{Intercept} \right)} (1)$$

Where $\hat{L}$ represents the maximized likelihood value from both our fitted model ($M_{Full})$and the null model ($M_{Intercept}$).

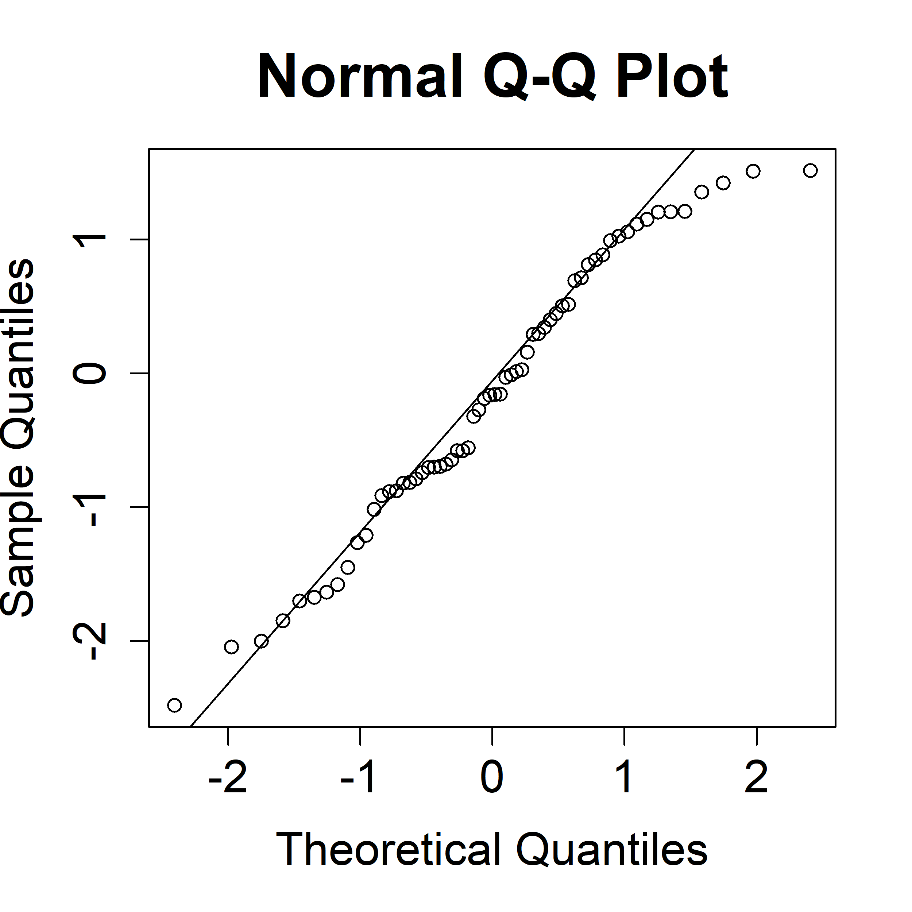

**Supplemental Figure 1:** Q-Q Plot to assess data distribution underlying our final negative binomial model
