## Supplementary material for "Quantifying the relationship between SARS-CoV-2 wastewater concentrations and building-level COVID-19 prevalence at an isolation residence using a passive sampling approach": Manuscript.docx(wouldn't properly convert to pdf)

Keywords: SARS-CoV-2; college campus monitoring; passive sampling; wastewater-based epidemiology; fecal shedding; COVID-19

Highlights

- Daily SARS-CoV-2 RNA loads in building-level wastewater were positively associated with the total number of COVID-19 positive individuals in the residence
- The variation in individual fecal shedding rates of SARS-CoV-2 extended four orders of magnitude
- Wastewater sample replicates were highly correlated using distinct processing pipelines in two independent laboratories
- While the isolation residence was occupied, SARS-CoV-2 RNA was detected in all passive samples

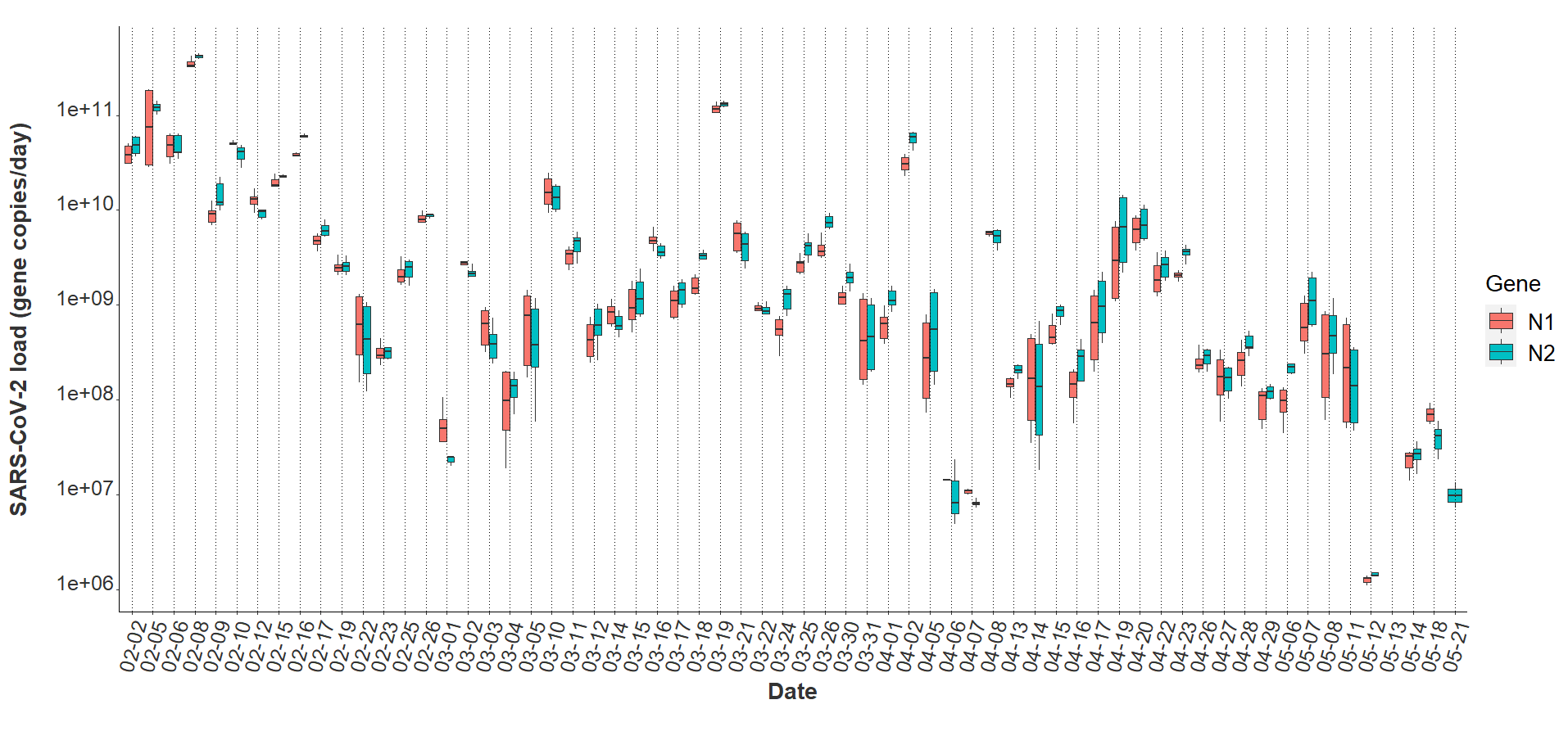

**Fig 1.** N1 and N2 daily viral loads from the COVID-19 isolation residence, February 1, 2021 – May 21, 2021 (n=64).

| **Table 1.** Quantification of viral loads and viral loads per population in the isolation residence | |
| --- | --- |
| **Characteristic** | **Value** |
| Viral gene copies per day |  |
| Median, N2 | 1.29 x 10^9^ |
| Range (min/max), N2 | 1.38 x 10^6^, 4.53 x 10^11^ |
| Median, N1 | 1.04 x 10^9^ |
| Range (min/max), N1 | 1.11 x 10^6^, 4.27 x 10^11^ |
| Viral gene copies per person per day |  |
| Median, N2 | 1.01 x 10^8^ |
| Range (min/max), N2 | 4.29 x 10^5^, 4.86 x 10^9^ |
| Median, N1 | 6.87 x 10^7^ |
| Range (min/max), N1 | 5.70 x 10^5^, 4.41 x 10^9^ |
| Water usage per person per day (L) |  |
| Median | 83.4 |
| Range (min/max) | 20.3, 479.7 |

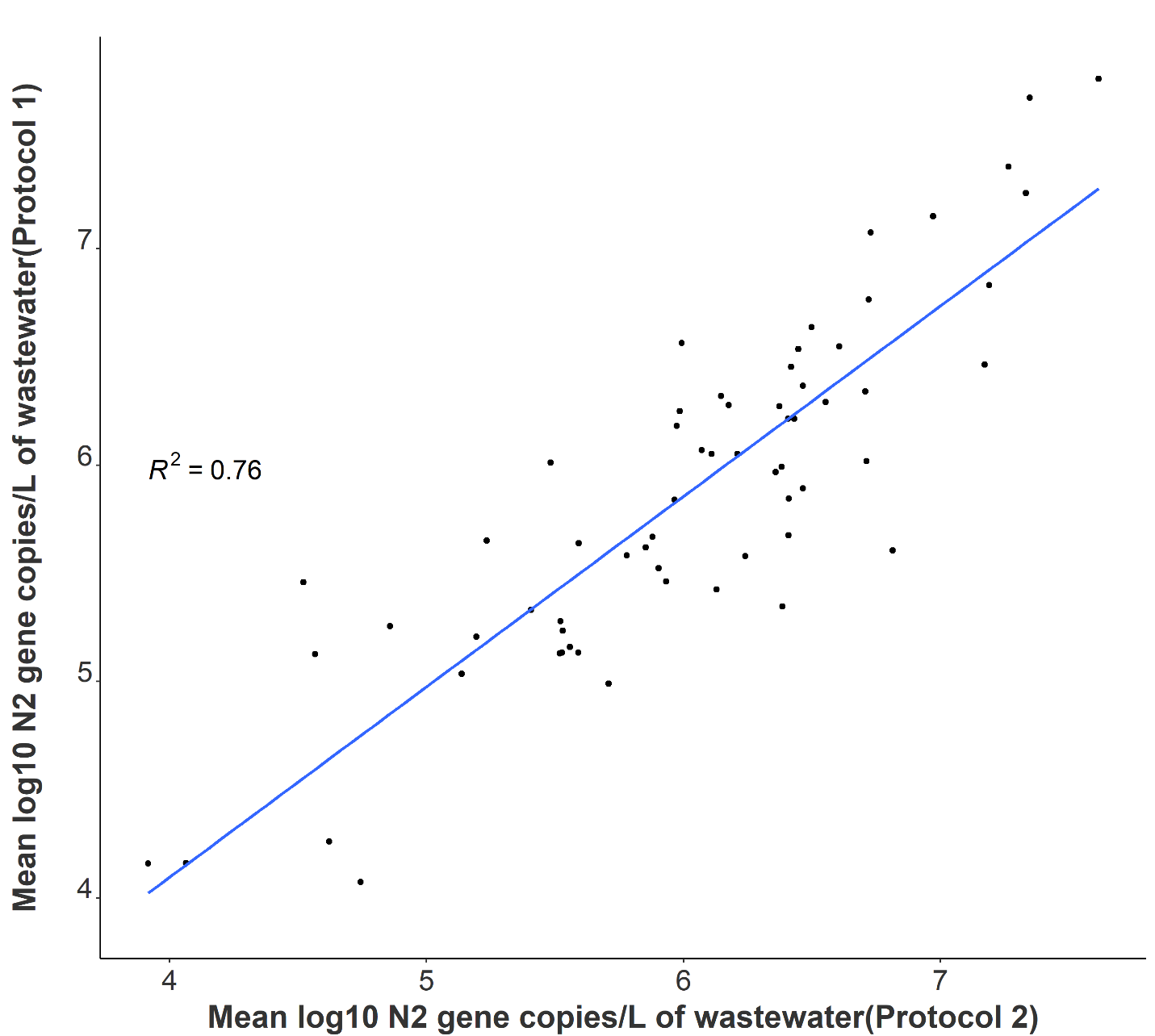

**Fig 2.** Correlation between independent average N2 log-transformed viral concentrations on identical samples utilizing separate processing and analysis pipelines (n = 64).

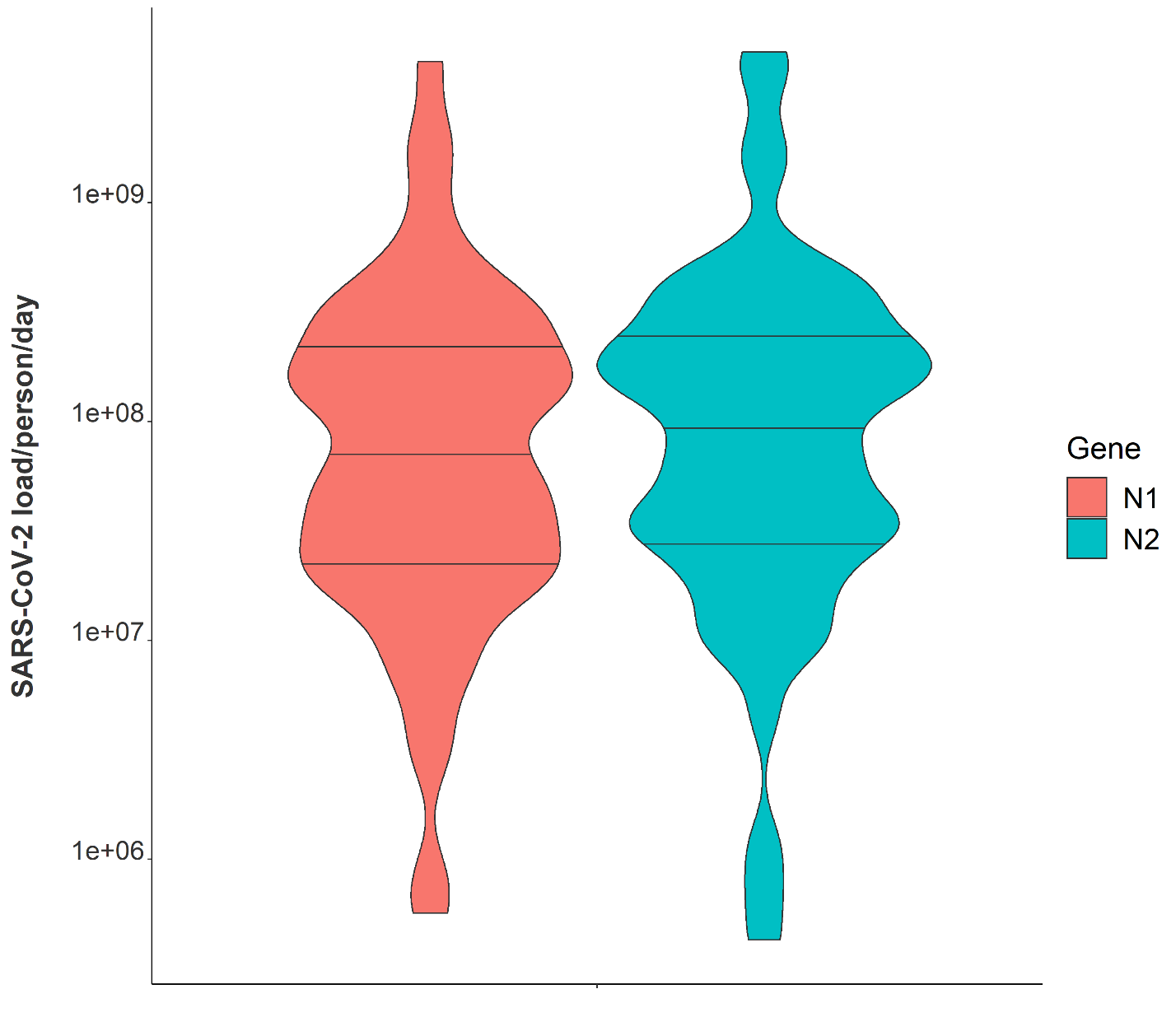

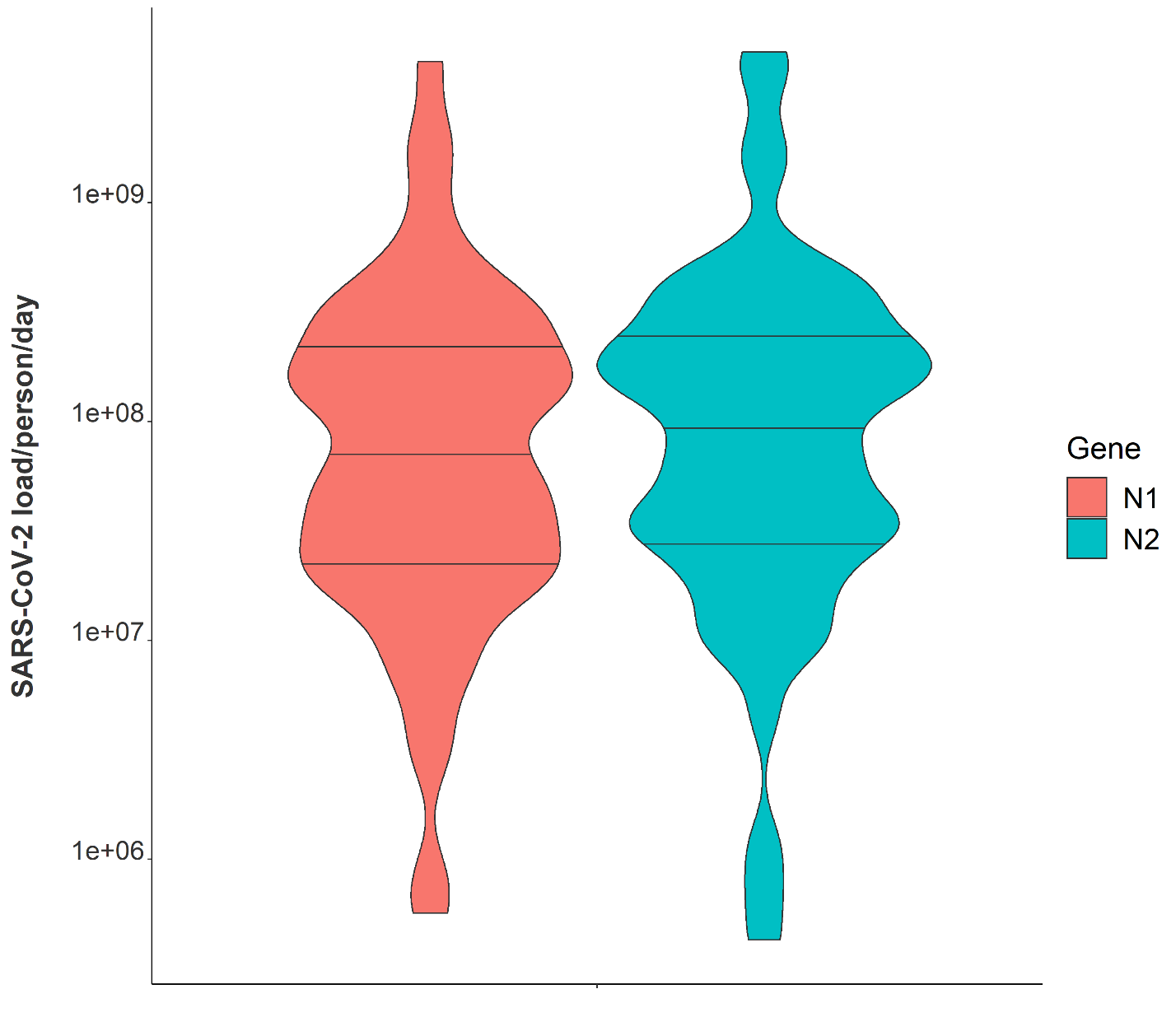
**Fig 3**. Violin plot showing distributions of N1 and N2 average daily wastewater viral loads per individual in isolation from February 1, 2021 – May 21, 2021. Note: Markers shown are median, 25^th^ and 75^th^ quantiles.

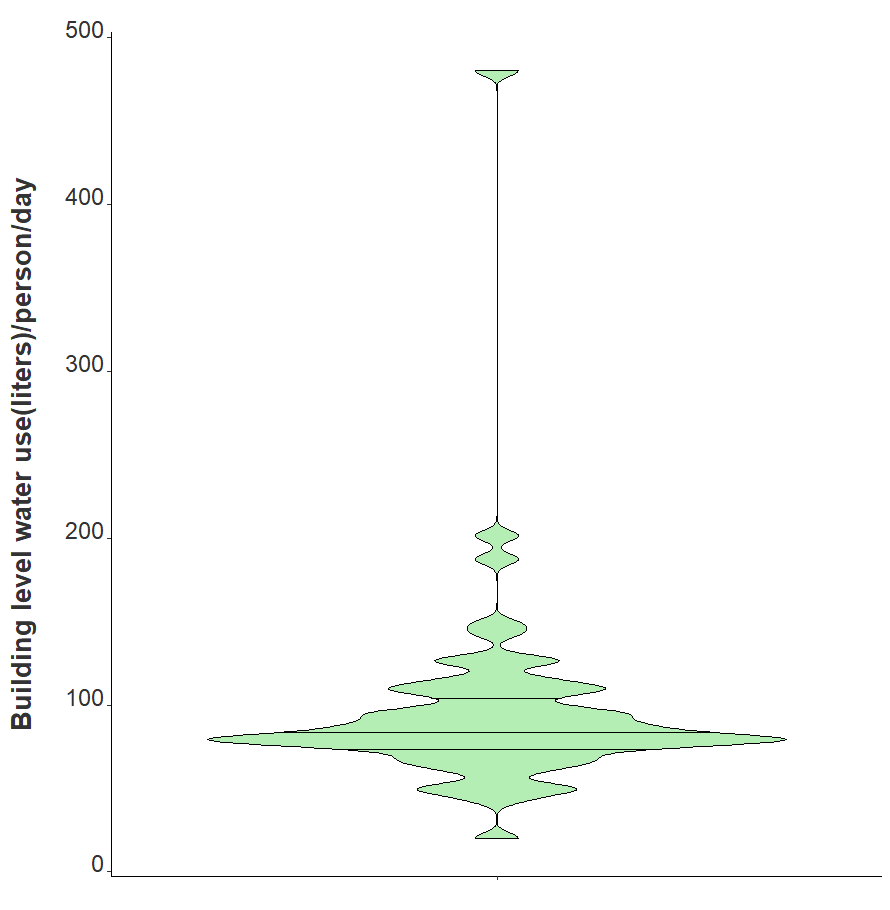

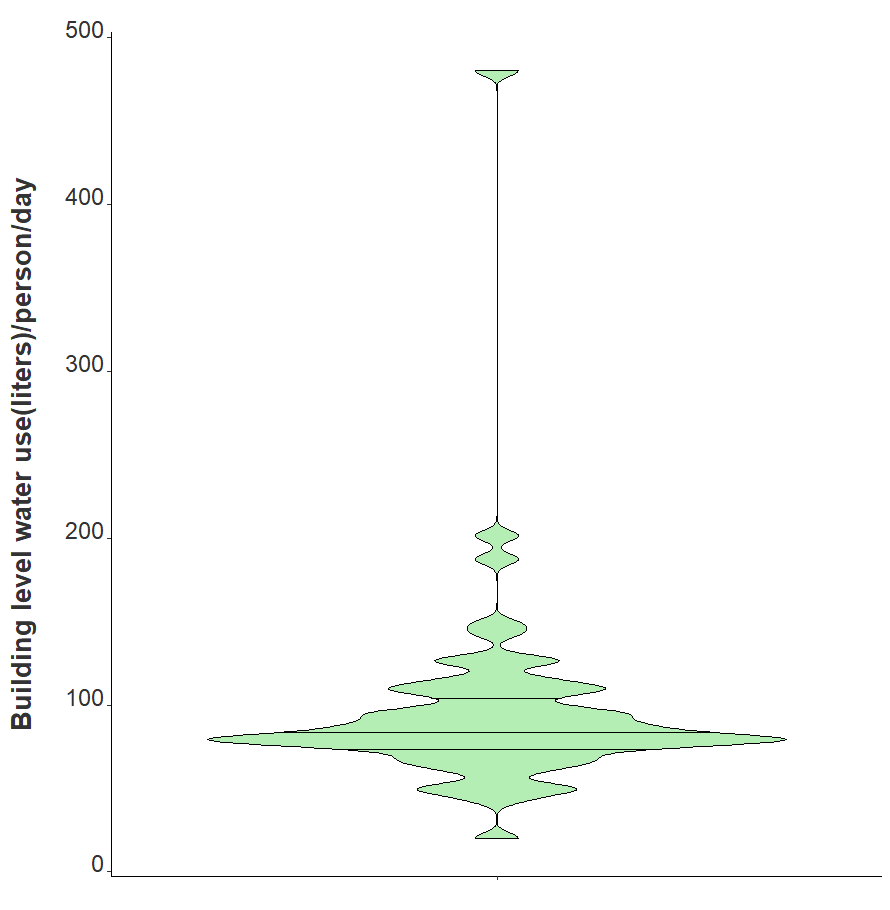
**Fig 4.** Violin plot showing distribution of average daily building-level water use per individual in isolation from February 1, 2021 – May 21, 2021. Note: Markers shown are median, 25^th^ and 75^th^ quantiles.

**Table 2.** Time series negative binomial models to quantify associations between captured viral loads and defined patient populations, Massachusetts 2021. Note: IRR = Incidence Rate Ratio.

| Factor | Univariate IRR | Univariate  p-value | Adjusted IRR | Adjusted  p-value |
| --- | --- | --- | --- | --- |
| BRSV Recovery | 0.99 | 0.40 | 0.99 | 0.02 |
| % Female Occupants | 1.05 | <0.001 | 1.05 | <0.001 |
| Log Viral Gene Copies | 2.08 | <0.001 | 1.47 | <0.001 |

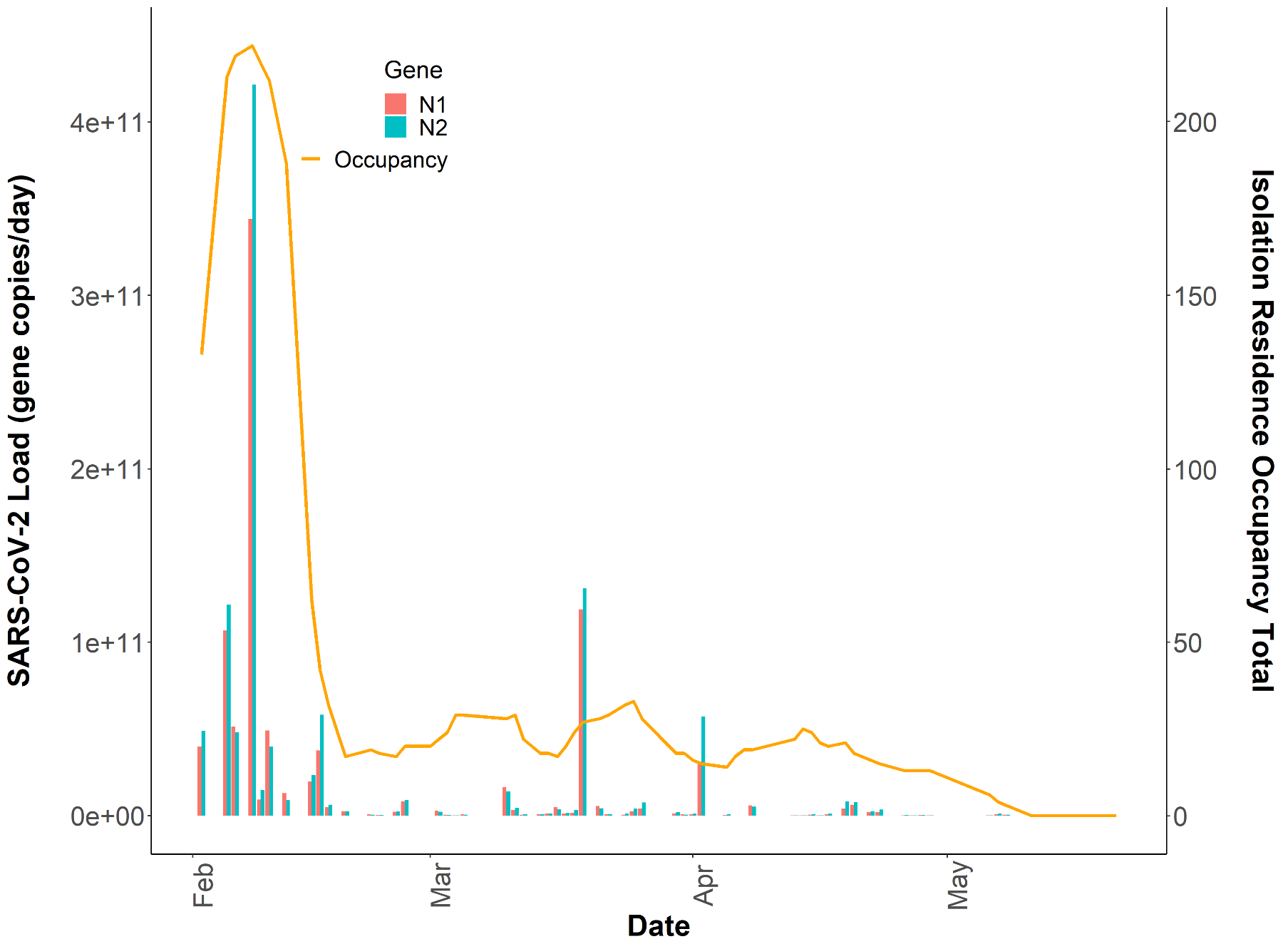

**Fig 5.** Total daily isolation building occupancy (line) plotted with N1 and N2 gene copies/day (bars) during the Spring 2021 academic semester**.** Note**:** Both SARS-CoV-2 daily wastewater viral loads and daily isolation residence occupancy totals are reported on linear y-axes (n=64).

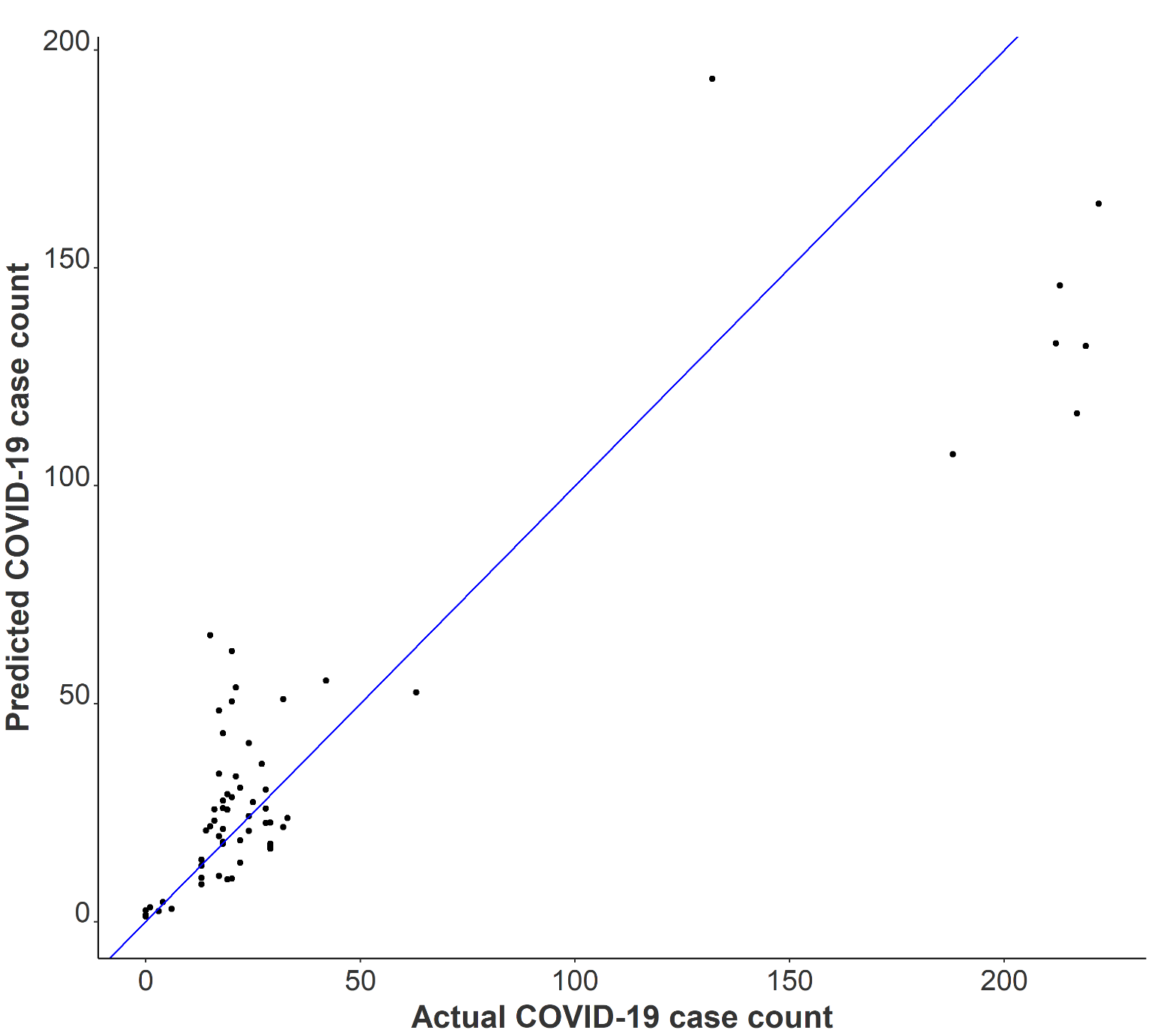

**Fig 6.** Observed COVID-19 isolation residence occupancy plotted against predicted COVID-19 isolation residence occupancy using negative binomial modeling with Spring 2021 data.
